# Sunbed users aged 18-24 views on sunbed regulation policies in the UK

**DOI:** 10.64898/2026.01.07.25342542

**Authors:** Angela M. Rodrigues, Lauren M. Hoult, Tracy Epton, Rachel Abbott, Paul Court

## Abstract

An online, cross-sectional survey was conducted to understand support for potential sunbed regulation strategies in the UK among 502 UK-based young adults (aged 18-24). Sunbed users showed strong support for sunbed regulation in the UK overall, particularly for staff supervision, mandatory health warnings, and business licensing, policies already implemented in some UK regions. Framing policies around public health protection rather than restricting personal freedom significantly enhanced support. These findings suggest a phased regulatory approach, starting with measures like stricter licensing and promotion of safer tanning alternatives that align with sunbed users’ sentiment, may build broader support for stronger regulation over time. This manuscript has been submitted to the British Journal of Dermatology and is currently under review.

## Dear Editor

Indoor tanning before age 35 increases melanoma risk by around 60%, and frequent users are 2–3 times more likely to develop the disease^1^. Despite age restrictions in the UK (a restriction on <18 years), sunbed use persists among minors and young adults, highlighting gaps in regulation compliance^2^. Internationally, approaches vary from complete bans to under-18 restrictions, but evidence suggests that stronger regulatory frameworks, combined with public health messaging, can reduce exposure and support skin cancer prevention^3^. Understanding public attitudes towards regulatory approaches could inform skin cancer prevention policy. We conducted a survey to assess support for potential sunbed regulation strategies in the UK.

An online, cross-sectional survey was conducted between February and June 2025 (https://osf.io/7xzmn/)^4^. Using purposive sampling, we recruited 502 UK-based young adults (aged 18–24; (M_age_ = 21.73, SD_age_ = 1.76) who had used sunbeds in the past year. Measures included demographics (age, ethnicity), social deprivation, skin cancer risk, sun protection behaviours, willingness to use alternatives, and policy support. Respondents rated support (1–5) for eight policies: (i) complete ban on commercial sunbed, (ii) locations bans (e.g., gyms, beauty salons, hotels, near schools), (iii) sunbed tax, (iv) removing VAT from safer tanning alternatives, (v) restricting sunbed advertising, especially on unlimited usage offers and health benefit claims, (vi) mandating health warning signs^1^, (vii) requiring staff supervision^1^, and (viii) businesses licensing system^2^. To determine if sunbed frequency use in the last year predicted each policy use, hierarchical linear regressions controlled for whether participants stopped using sunbeds, ethnicity (dummy coded with White as reference vs Black, Asian, and Mixed), school type (state vs independent), and free school meal eligibility. A MANOVA determined if support varied by sunbed use (stopped vs not), controlling for demographics. Ethical approval was obtained from Northumbria University (ref: 9369), and informed consent was obtained from all participants.

Support was highest for policies already implemented in some UK regions: staff supervision (net support = 75.5%), mandating health warnings (74.9%), and business licensing (73.1%). Other supported policies included removing VAT from safer tanning options (44.3%) and restricting advertising (38.2%). Views were mixed on location bans (0.6%). Policies mostly opposed were total commercial bans (−6.7%) and sunbed tax (−22.5%) (Figure 1).

**Figure 1:**
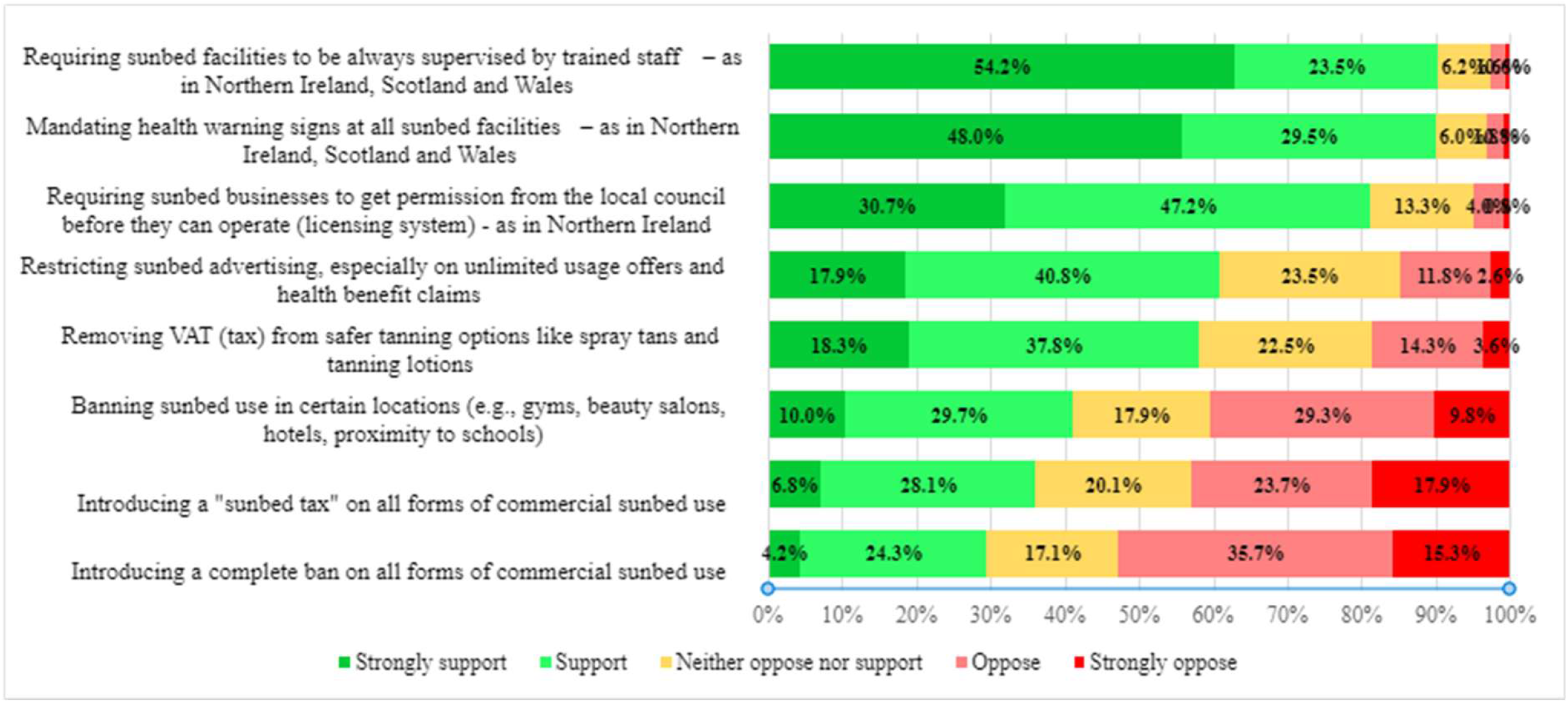
Sunbed users aged 18-24 support for eight sunbed policies in the United Kingdom.

Hierarchical linear regressions showed final models were significant for most policies (Δ*R*^*2*^ =.04-.20, all *p* ≤.025) after controlling for demographics, except business licensing. More frequent sunbed use predicted lower support for total bans, location bans, sunbed tax, removing VAT from alternatives, restricting advertising, and mandating health warnings (*β* = -.08-.20, all p ≤.031).

Multivariate analyses revealed significant effects of sunbed use, ethnicity, and free school meal eligibility (Pillai’s Trace =.039-.093, all *p* ≤.042). Former users showed consistently higher support than current users across most policies (*F* = 6.61-19.56, all *p* ≤.011), except mandating health warnings, staff supervision, and business licensing. Free school meal-eligible participants showed greater support for removing VAT from alternatives and restricting advertising (*F* = 4.84-12.39, *p* ≤.001). Support varied by ethnicity for location bans, sunbed tax, mandating health warnings, and staff supervision (*F* = 2.77-4.81, *p* ≤.042); post-hocs showed Asian participants reported lower support than White and Black participants, while Mixed ethnicity participants showed greater support than White and Black participants for some policies.

Sunbed users showed strong support for sunbed regulation in the UK overall, particularly for staff supervision, mandatory health warnings, and business licensing, policies already implemented in some UK regions. Support varied by frequency of sunbed use, as well as ethnicity, and socioeconomic background, highlighting the need for tailored policy approaches. Framing policies around public health protection rather than restricting personal freedom significantly enhanced support, echoing findings from smoking^5^ and net zero policy research^6^. These findings suggest a phased regulatory approach, starting with measures like stricter licensing and promotion of safer tanning alternatives that align with sunbed users’ sentiment, may build broader support for stronger regulation over time. Effective policy development should incorporate user perspectives and emphasise diverse benefits (health, youth safety, economic, industry standards) to gain broader acceptance than health-risk messaging alone^7^.

Limitations include self-report bias and potential sampling error, though the large, demographically balanced sample of sunbed users increases confidence in findings. Future research should explore whether these patterns extend to the general public.

With rising melanoma incidence, strengthening evidence-based sunbed policy and public awareness of risks is urgent.

## Data Availability

All data produced are available online at Open Science Framework.

https://osf.io/7xzmn/

## Acknowledgments

We would like to thank all the participants who took the time to participate in this research, and the PPIE contributors for supporting this project throughout. We are grateful to Melanoma Focus for funding the study and helping with recruitment.

AR, TE, RA and PC developed the initial study design and secured funding for the study. LH and AR conducted preparation of study materials, data collection and analysis. AR and LH drafted the manuscript and its revised iterations. LH, AR, TE, RA and PC contributed and provided comments on data analysis and interpretation, and report drafts. All co-authors have reviewed and agreed the final draft of the paper submitted for publication. This study was funded by Melanoma Focus (Registered Charity No. 1124716). This analysis uses a subset of data from a survey on sunbed use and sunbed alternatives^4^.

## Footnotes

1 As in Northern Ireland, Scotland and Wales

2 As in Northern Ireland

## References

1. Boniol M, Autier P, Boyle P and Gandini S. Cutaneous melanoma attributable to sunbed use: systematic review and meta-analysis. BMJ: British Medical Journal 2012; 345: e4757. DOI: 10.1136/bmj.e4757.

2. Melanoma Focus. https://melanomafocus.org/news-blog/28-of-uk-adults-are-using-sunbeds-as-skin-cancer-rates-rise/ (2025, 2025).

3. Long A, O’Malley S and Collins S. The sunbed trend in Ireland. British Journal of Dermatology 2024; 191: 630–631. DOI: 10.1093/bjd/ljae209.

4. Hoult L, Rodrigues, A., Epton, T., Abbott, R., Court, P. Exploring young adults’ perspectives on alternatives to sunbed use: a cross-sectional survey., osf.io/7xzmn (2025).

5. Kock L, Shahab L, Moore G, et al. Assessing the profile of support for potential tobacco control policies targeting availability in Great Britain: a cross-sectional population survey. Tobacco Control 2024; 33: 221. DOI: 10.1136/tc-2022-057508.

6. Poortinga W, Whitmarsh L, Steentjes K, et al. Factors and framing effects in support for net zero policies in the United Kingdom. Frontiers in Psychology 2023; Volume 14 - 2023. Original Research. DOI: 10.3389/fpsyg.2023.1287188.

7. Mantzari E, Reynolds JP, Jebb SA, et al. Public support for policies to improve population and planetary health: A population-based online experiment assessing impact of communicating evidence of multiple versus single benefits. Social Science & Medicine 2022; 296: 114726. DOI: 10.1016/j.socscimed.2022.114726.

